# Contextual Barriers and Facilitators Influencing Implementation Fidelity of School-Based Preventive Chemotherapy for Schistosomiasis: A Qualitative Study in Two Endemic Districts in the Central Region, Ghana

**DOI:** 10.64898/2026.04.27.26351652

**Authors:** Haruna Moshi, Betwel J. Msugupakulya, Mahnaz Vahedi, Franklin N. Glozah

## Abstract

**Background:** Schistosomiasis remains a significant neglected tropical disease of public health concern, particularly in Sub-Saharan African countries, including Ghana. For decades, school-based preventive chemotherapy (PC) has been the mainstay of schistosomiasis elimination in Ghana; however, implementation fidelity across districts falls below WHO recommendations, leading to persistent transmission. This study explores contextual factors that influence the implementation fidelity of school-based preventive chemotherapy for schistosomiasis in endemic districts.

**Methods:** A phenomenological qualitative study design using a maximum-variation purposive sampling technique was conducted in two endemic districts (Gomoa East and Awutu Senya East) in Ghana, with 21 participants, six key informant interviews with district Neglected Tropical Diseases (NTDs) coordinators and School Health Education Program (SHEP) coordinators, and 15 in-depth interviews with head teachers and SHEP teachers. Recruitment of participants and data collection were conducted from 20/06/2025 to 30/07/2025 using semi-structured interview techniques and were thematically analyzed in NVivo version 15, guided by Braun & Clarke (2006). The thematic analysis blended inductive and deductive coding techniques; inductive allowed themes to emerge from the data, while deductive was guided by Damschroder et al. (2022) Consolidated Framework for Implementation Research (CFIR), and Carroll et al. (2007), a framework for implementation fidelity.

**Results:** Implementation fidelity was shaped by the interplay of sociocultural, organizational, and health system factors. Key barriers included socio-cultural beliefs and perceptions on treatment uptake, drug-related fears and adverse reactions, logistical constraints and resource limitations, lack of training and incentives for frontline implementers, inadequate community sensitization and engagement, and gaps in coverage and adherence to program protocols. Conversely, fidelity was much better in situations where awareness had been raised, the community was effectively engaged, frontline implementers were motivated, and collaboration was strong among the health and education sectors, suggesting that high fidelity can be achieved through a systemic response.

**Conclusion:** Implementation fidelity of school-based preventive chemotherapy is a context-dependent, system-driven process shaped by the complex interaction of socio-cultural and structural factors. Moving forward, to enhance fidelity and achieve sustained schistosomiasis control requires a shift toward a community-centered delivery approach that emphasizes community sensitization and engagement, reliable logistical support, effective training and motivation for frontline implementers, and intersectoral collaboration.

## Background

Schistosomiasis is a neglected tropical disease caused by parasitic trematodes of the genus Schistosoma. It remains a significant public health problem in many low- and middle-income countries, particularly in sub-Saharan Africa. In Africa, Schistosoma mansoni and Schistosoma haematobium are the predominant species, responsible for intestinal and urogenital schistosomiasis, respectively (4). Transmission occurs through contact with freshwater contaminated with cercariae released by infected freshwater snails, which penetrate human skin and mature into adult worms within the host (5). Because transmission is strongly linked to routine water contact and inadequate water, sanitation, and hygiene infrastructure, schistosomiasis persists in communities with frequent exposure and limited environmental control. These characteristics make preventive chemotherapy (PC) delivered at schools and the community level a cornerstone of schistosomiasis control strategies (6).

Globally, schistosomiasis affects over 250 million people across 78 endemic countries, with more than 90% of the disease burden concentrated in sub-Saharan Africa (5). School-aged children bear a disproportionate share of this burden because they frequently engage in activities that increase water contact, such as swimming and bathing in contaminated freshwater bodies, making them a primary target for school-based preventive chemotherapy programs (7). In Ghana, schistosomiasis remains endemic across all regions, with persistent transmission reported in several coastal and peri-coastal districts, including Gomoa East and Awutu Senya East in the Central Region, where school-based preventive chemotherapy has been implemented for several years (Kwabi et al., 2024). In Gomoa East specifically, the current study indicates an upsurge in schistosomiasis prevalence from 10.4% in 2021 to 15.5% in 2023 (9).

Ghana has been implementing school-based preventive chemotherapy since 2008, led by the Ghana Health Service (GHS) in collaboration with the Ghana Education Service (GES) as part of the national strategy for controlling neglected tropical diseases (10). The program is implemented through the School Health Education Program (SHEP), where trained teachers administer praziquantel to eligible school-aged children during mass drug administration campaigns (11). Evidence from Ghana Health Service reports, and recent studies shows that although the program has been implemented across all endemic districts, treatment coverage has not consistently reached the WHO-recommended threshold of at least 75% of school-aged children (10). For example, recent campaign data from the Central Region indicate that coverage in some districts, including Gomoa East and Awutu Senya East, has fluctuated between approximately 68% and 74.2%, suggesting that a proportion of the target population remains untreated during some campaign cycles (12). Adherence to program protocols has been inconsistent across districts, with the continued prevalence of schistosomiasis among school-aged children (13). This indicates that the program has not yet achieved the desired level of effectiveness in interrupting transmission, which is due to the presence of contextual and operational challenges in program delivery.

To better understand these challenges, this study was guided by Damschroder et al. (2022) Consolidated Framework for Implementation Research (CFIR) and the Carroll et al. (2007) implementation fidelity framework. The CFIR provides a comprehensive lens for examining multilevel determinants of implementation, including intervention characteristics, the outer setting, the inner setting, individual characteristics, and implementation processes (2). In this study, the outer setting focuses on sociocultural beliefs, community perceptions, and social influences that shape acceptance of preventive chemotherapy. The inner setting examines organizational capacity, support, and the overall implementation climate within schools and district health systems. The characteristics of individuals consider the knowledge, beliefs, and motivation of frontline implementers. The implementation process domain focuses on how the intervention is planned and executed within schools. To complement CFIR, the implementation fidelity framework was used to assess the extent to which the program is delivered as intended, including key dimensions such as coverage, adherence to protocols, quality of delivery, and timing (3). These frameworks provide a structured and integrated approach for examining how contextual factors influence both the delivery and effectiveness of school-based preventive chemotherapy.

Evidence suggests that, beyond drug availability, the effectiveness of preventive chemotherapy depends heavily on implementation fidelity (the degree to which program components are delivered as intended in terms of coverage, timing, delivery processes, and adherence to guidelines) (14,15). Persistent fidelity gaps, therefore, represent a critical but underexplored driver of suboptimal program outcomes in these settings. This study aimed to explore contextual barriers and facilitators influencing the implementation fidelity of school-based preventive chemotherapy for schistosomiasis in the Gomoa East and Awutu Senya East districts of the Central Region of Ghana.

## Methods

### Study Design

A phenomenological qualitative study design was employed to explore the lived experiences of stakeholders involved in implementing school-based preventive chemotherapy for schistosomiasis. This design was selected to enable in-depth exploration of contextual, organizational, and individual factors affecting the implementation process within their specific district contexts, thereby aligning with the study’s objective. The design was guided by two complementary frameworks: the Consolidated Framework for Implementation Research (CFIR), which provided a lens to examine multi-level contextual determinants (2), and Carroll et al’s Implementation Fidelity Framework, which conceptualizes how these determinants affect program adherence, coverage, frequency, and quality (3).

### Study setting

This study was conducted in two schistosomiasis-endemic districts in the Central region (Gomoa East District and Awutu Senya East) (10). These districts are characterized by tropical climates and diverse water bodies, including the Pra and Akrudu rivers and the Gulf of Guinea, which provide an ideal environment for freshwater snails and favor schistosomiasis transmission, given the community’s dependence on these water sources. These districts have implemented school-based preventive chemotherapy programs for several years, providing an established context to examine how multi-level factors influence fidelity. Health service delivery includes district hospitals, polyclinics, health centers, and community-based health planning and services compounds. At the same time, education is delivered through a mix of public and private basic schools, which serve as the primary platforms for mass drug administration.

### Study population and Sampling

The study population comprised districts and school-level implementers involved in planning, coordinating, delivering, and supervising preventive chemotherapy. Participants were purposively selected to include respondents with experience in program implementation. Maximum variation sampling was used to capture perspectives across different implementation levels, environments, and roles. A total of 21 participants were interviewed, including four district Neglected Tropical Diseases (NTDs) program coordinators, two district School Health Education Program (SHEP) coordinators, nine School Health Education Program (SHEP) teachers, and six head teachers. The sample size was determined based on thematic saturation, the point at which additional interviews yielded no new themes relevant to the study objectives.

### Inclusion and exclusion criteria

The study included participants residing in the Awutu Senya and Gomoa East districts who have been active implementers of school-based preventive chemotherapy programs targeting schistosomiasis for at least six months. Participants who refused to participate by not signing the informed consent form were excluded from the study.

### Data collection techniques

Recruitment of participants and data collection started on 20/06/2025 and ended on 30/07/2025, using semi-structured interviews. This method was selected to provide the researcher with flexibility in guided conversations, enabling in-depth exploration of participants’ lived experiences while ensuring that key implementation-fidelity topics were consistently addressed across interviews. A total of 21 interviews were conducted face-to-face in private locations within participants’ workplaces to maintain confidentiality and minimize interruptions. Six key informant interviews with district Neglected Tropical Diseases (NTDs) program coordinators and district School Health Education Program (SHEP) coordinators. The remaining 15 were in-depth interviews with SHEP teachers and head teachers. Each interview lasted approximately 30 to 60 minutes and was audio recorded with participants’ consent. Two separate interview guides were prepared, one for key informant interviews and another for in-depth interviews. Each guide consisted of four main sections. The first section gathered background information on participants, including their roles and experience in program implementation. The second section examined perceived barriers and facilitators affecting the program’s implementation fidelity. The third section focused on fidelity components (coverage, content delivered, duration, and frequency of the program). The final section asked about recommended strategies for enhancing program implementation fidelity. The guides mainly consisted of open-ended questions, supplemented by follow-up probes to encourage participants to elaborate based on their experiences.

### Data analysis

All audio-recorded interviews were transcribed verbatim by listening to the recordings and typing the content into Microsoft Word documents. Where interviews were conducted in the local language, they were first transcribed in the original language and subsequently translated into English to ensure accuracy of meaning. The transcripts were reviewed alongside the audio recordings to ensure completeness and correctness. After transcription, the Word documents were imported into NVivo software (version 15) to facilitate systematic coding, organization of data, and thematic analysis. Data were analyzed thematically, guided by Braun and Clarke’s six-step framework, which involved familiarization with the data, generating initial codes, developing potential themes, reviewing and refining themes, and production of an analytical narrative (1). Both inductive and deductive coding approaches were applied: inductive coding allowed new themes to emerge from participant narratives, while Damschroder et al. (2022) Consolidated Framework for Implementation Research (CFIR), and Carroll et al. (2007) Implementation Fidelity Framework guided deductive coding. To enhance analytical rigor, inter-coder reliability techniques were used where two qualitative analysts independently coded a subset of transcripts and compared results to ensure reliability and consistency.

### Ethical Considerations

Ethics approval for this study was obtained from the Ghana Health Service Ethics Review Committee (Approval No: GHS-ERC 053/05/25). Administrative permissions were also secured from the relevant district health and education directorates before data collection. All participants were provided with an information sheet detailing the study’s purpose, procedures, potential risks, and benefits. Written informed consent was obtained from each participant as proof of voluntary participation. Participants were assured of confidentiality, anonymity, and their right to refuse or withdraw from the study at any point without any consequences or loss of benefits. Numerical codes were used in place of personal identifiers to ensure anonymity; all interviews were conducted in private, undisclosed locations, and data were securely stored with a password. No financial incentives were given to participants to minimize the potential for undue influence on participants or their responses.

### Trustworthiness of the study

Methodological rigor was ensured through adherence to Lincoln and Guba’s (1985) criteria of trustworthiness: credibility was enhanced through triangulation and member checking; dependability and confirmability were supported through audit trails, reflexive journaling, and peer debriefing; and transferability was promoted through detailed contextual descriptions to enable comparison with similar schistosomiasis-endemic settings.

## RESULTS

Analysis generated several interconnected themes that collectively illustrate the contextual factors influencing implementation fidelity of the school-based preventive chemotherapy (PC) program, as well as factors directly related to the intervention’s structure and delivery.

### Socio-demographic characteristics of study participants

A total of 21 participants were included, comprising District NTD coordinators, SHEP coordinators, SHEP teachers, and head teachers from Gomoa East and Awutu Senya East. The sample was nearly balanced by sex (11 females and 10 males), with ages ranging from 21 to 55 years. District-level coordinators tended to be older, while teachers were generally younger, particularly among SHEP teachers.

Participants reported between 2 and 13 years of professional experience. District NTD and SHEP coordinators were more experienced, whereas SHEP teachers reflected a mix of early and mid-career professionals. Head teachers fell within a moderate to higher experience range. Participants were drawn from both districts, with slightly higher representation from Gomoa East. Detailed socio-demographic characteristics of the participants are presented in Table 1.

**Table 1:** Socio-demographic characteristics of study participants (N = 21)

| Participant type | Sex | Age range | Years of experience |
| --- | --- | --- | --- |
| **District NTDs Coordinator (KII)** |  |  |  |
| NTD Program Coordinator 1 | Male | 41-45 | 8-10 Years |
| NTD Program Coordinator 2 | Male | 46-50 | 4-7 Years |
| NTD Program Coordinator 3 | Female | 36-40 | 4-7 Years |
| NTD Program Coordinator 4 | Male | 41-45 | 8-10 Years |
| **District SHEP Coordinators (KII)** |  |  |  |
| SHEP Coordinator 1 | Female | 36-40 | 4-7 Years |
| SHEP Coordinator 2 | Male | 51-55 | 14-17 Years |
| **SHEP Teachers (IDI)** |  |  |  |
| SHEP Teacher 1 | Female | 26-30 | 1-3 years |
| SHEP Teacher 2 | Female | 36-40 | 4-7 years |
| SHEP Teacher 3 | Male | 31-35 | 4-7 years |
| SHEP Teacher 4 | Female | 26-30 | 1-4 years |
| SHEP Teacher 5 | Male | 41-45 | 8-10 Years |
| SHEP teacher 6 | Male | 36-40 | 4-7 years |
| SHEP Teacher 7 | Female | 41-45 | 8-10 Years |
| SHEP Teacher 8 | Female | 26-30 | 4-7 Years |
| SHEP Teacher 9 | Male | 46-50 | 11-13 years |
| **Head Teachers (IDI)** |  |  |  |
| Head Teacher 1 | Female | 51-55 | 11-13 Years |
| Head Teacher 2 | Male | 46-50 | 8-10 Years |
| Head Teacher 3 | Male | 36-40 | 4-7 Years |
| Head Teacher 4 | Female | 46-50 | 8-10 Years |
| Head Teacher 5 | Female | 41-45 | 4-7 Years |
| Head Teacher 6 | Female | 41-45 | 4-7 Years |

### Barriers to implementation fidelity

Analysis identified multiple barriers to implementation fidelity operating across community, school, and health system levels. Key barriers included: (1) Socio-cultural beliefs and misconceptions on treatment uptake, (2) Drug-related fears and adverse reactions, (3) logistical constraints and resource limitations, (4) lack of training and incentives for frontline implementers, (5) inadequate community sensitization and engagement, and (6) gaps in coverage and adherence to program protocols. These barriers were mapped to CFIR outer and inner setting domains and to Carroll et al’s fidelity constructs (coverage, adherence, dose delivered, quality of delivery), illustrating how contextual and structural factors influence implementation outcomes. A detailed breakdown of themes identified from the data is presented in Table 2.

**Table 2:** Key themes identified as barriers in implementing school-based preventive chemotherapy.

| Main Themes | Sub-Themes |
| --- | --- |
| 1. Socio-cultural beliefs and misconceptions on treatment uptake | • Cultural and religious beliefs<br>• Perception and myths surrounding the school-based preventive chemotherapy.<br>• Parental resistance to school-based preventive chemotherapy. |
| 2. Drug-related fears and adverse reactions | • Real drug-side effects<br>• Large size of the praziquantel drug<br>• Challenge in managing side effects<br>• Psychosomatic reaction and peer imitation |
| 3. Logistical constraints and resource limitations | • Insufficiency of measuring tools<br>• Inadequate transportation logistics |
| 4. Lack of training and incentives for frontline implementers | • Inadequate teachers' training<br>• Lack of incentives for frontline implementers<br>• Shortage of well-trained implementers |
| 5. Inadequate community sensitization and engagement | • Inadequate pre-MDA community sensitization<br>• Poor community engagement<br>• Insufficient awareness programs |
| 6. Insufficient program funds | • Inadequate program funding<br>• Delays in funding release<br>• Competing school-based programs |

### Socio-cultural beliefs and misconceptions on treatment uptake

Socio-cultural beliefs and misconceptions were frequently reported as the main barriers affecting program fidelity. Rumors that the praziquantel drug causes infertility or is harmful fueled mistrust, which triggered resistance among parents and children, leading to low intervention uptake, directly affecting fidelity dimensions of coverage and dose adherence. As highlighted by the district NTD coordinator and SHEP Teacher:

> “The big problem here is that people think the drugs we give out for free are poison and cause infertility… These are super drugs, and they have something to do with controlling birth…. That is their perception”. (KII, NTD Program Coordinator 1).
>
> “There are a lot of rumors in this community, some people believe the drug causes infertility…, Today it’s worms, tomorrow it is infertility” (IDI, SHEP Teacher 2).

These historical myths and rumors surrounding school-based preventive chemotherapy triggered parents to forcibly remove their children and stop them from going to school during the mass drug administration day. One respondent said that:

> “You see parents picking their kids up early, some telling me not to give my child anything…, Some don’t come to school at all on the day of administration because their parents tell them not to go. It’s becoming a trend.” (IDI, Head Teacher 2).

Some cultural and religious practices restricted participation in school-based preventive chemotherapy, especially when trusted faith leaders advised against drug administration, framing it as a harmful or satanic strategy to weaken children. One respondent shared that:

> “I remember one pastor advised students not to take any medication given in schools, he called it ‘Satan’s strategy to weaken children’” (IDI, Head Teacher 1).

These beliefs further affected the health-seeking behavior of the community when someone got sick with schistosomiasis, especially in rural and conservative areas, where sick children with schistosomiasis or drug reactions are referred to traditional healers, prayer camps, or herbal medicine over formal health systems. As revealed by one teacher who stated that:

> “One parent told me openly: ‘If my children feel sick, I won’t bring them to a hospital. The pastor will pray for them, or we’ll use herbs at home.” (IDI, SHEP Teacher 3).

This narrative was strongly supported by another respondent, who is a neglected tropical diseases program coordinator:

> “If a child shows side effects, some parents do not go with that child to the health facility. They rush them to an herbalist or prayer camp because they believe the medicine we give is ‘foreign’ and harmful, but local herbs are safe.” (KII, NTD Program Coordinator 1).

These opinions serve as evidence of the current trust gap existing between biomedical and traditional/faith-based interventions. Cultural and religious systems negatively influenced the use of biomedical health care systems, which resulted in low community responsiveness to school-based preventive chemotherapy programs. Such narratives show how misinformation and negative beliefs impede the implementation fidelity of the school-based preventive chemotherapy program

### Drug-related fears and adverse reactions

Fear of side effects emerged as a significant determinant of both parental resistance and pupil reluctance. Participants commonly mentioned dizziness, nausea, vomiting, stomach pain, and drowsiness as reactions pupils experienced after taking praziquantel, especially when children had not eaten before drug administration. One respondent said:

> “One girl fainted; we found out later she hadn’t eaten since that morning. Some students experienced heavier bleeding than usual during their menstrual cycles after taking the drug. It’s very prevalent, particularly in schools [that do] not offer a feeding program.” (IDI, SHEP Teacher 5).

The standard protocol is to have children eat before they take the drug, but that isn’t always possible. While side effects were revealed among pupils, others were described as psychosomatic responses triggered by fear. One respondent narrated that:

> “I went to one school, I found almost 50 pupils lying down that they are reacting to the drug, but nobody has given the drugs…, once one starts complaining, others panic and say they feel sick.” (KII, NTD Program Coordinator 1).

Fear related to the size of the praziquantel tablets also affected uptake. Many pupils struggled to swallow the large tablets, and some spat them out or pretended to swallow them while discarding the tablets when teachers were not watching. Participants said:

> “The tablet is too big and strong, students get scared and run away or pretend as if they took it. When no one is observing, they will discard it,” (IDI, SHEP Teacher 4).

This statement was strongly supported by another respondent, who is a head teacher in one of the schools, who added that:

> “You know, in the last MDA, students at my school were asking, madam, this time is not that white one (praziquantel)? That one is too big. I am unable to swallow it” (IDI, Head Teacher 3).

The control of adverse drug side effects added more worries and pressure to teachers and coordinators who are not well-trained on how to handle the clinical response. Respondents said that:

> “In many places, teachers are afraid to give this medicine to children because they don’t understand how to manage side effects and reactions of the drug…they are afraid to be blamed by parents, and pay the hospital bills” (KII, NTD Program Coordinator 2).

Drug side effects, psychosomatic reactions, and large tablet size created fear that contributed to low school-based preventive chemotherapy program uptake, which increased deviations from expected implementation fidelity standards, particularly in relation to coverage, dose, and content adherence in subsequent rounds.

### Logistical constraints and resource limitations

Participants highlighted several logistical challenges that hindered the smooth implementation of the school-based preventive chemotherapy program. Although praziquantel drugs were available in large quantities, all districts reported receiving inadequate supporting tools for implementing the school-based preventive chemotherapy. Many schools lacked adequate height-measuring poles or tapes, which forced teachers to improvise by marking walls to estimate children’s heights, which compromised precision, resulting in dosing inaccuracies. Respondents said:

> “Our district received only 80 measuring tapes for more than 300 schools. We distributed them…, and the rest of the school had to draw the wall, of course not accurate, but it is better than nothing” (KII, NTD Program Coordinator 1).
>
> “We had to make height charts on the wall in the classroom with chalk and approximate dosages by comparison. It’s not like we do it perfectly… What other options do we have?” (IDI, Head Teacher 2).

Problems of transportation were also cited in both districts. Transportation difficulties affected the timely distribution of drugs and supervision across schools. Allocation of vehicles was often limited, and fuel shortages delayed activities. Schools located in hard-to-reach communities frequently received drugs late or missed supervisory visits entirely. One respondent said that:

> “We don’t have vehicles, and nobody gives us money to go collect the drugs. We use our own money with no refund; it costs us” (IDI, SHEP Teacher 1).
>
> “Some schools are so far…, we are supposed to supervise all schools, but with only one vehicle, it is impossible” (KII, NTD Coordinator 1).

Delayed release of program funds was also highlighted. Participants described situations where money for community sensitization, fuel, or allowances was released a few days before the mass drug administration exercise began, limiting their ability to conduct preparatory activities, which weakened the fidelity of the implementation process, particularly planning and adherence to scheduled timelines.

> “There’s always a late release of funds… so all activities are usually done a few days before the program begins in a rush” (KII, NTD Program Coordinator 1).

Although the drugs themselves were largely in supply, logistical and resource limitations weakened the fidelity of the implementation process, particularly in relation to adequate planning and adherence to scheduled timelines.

### Lack of training and incentives for frontline implementers

Despite the central role played by teachers in implementing school-based preventive chemotherapy programs, no systematic financial or non-financial motivations are given to them; as a result, they become less committed to the program or are demotivated. One respondent said that:

> “Teachers are not given anything as rewards for the work. So, they say, why should I let parents come and insult me while I’m not getting anything… it really impacts their commitment to this program.” (KII, NTD Program Coordinator 1).

Lack of training for the teachers added a challenge to their ability to administer drugs and communicate effectively with parents about the program. Training was provided for only one teacher per school, and that training was conducted in a rush just a day before MDA, which limited their ability to master protocols. Respondents said:

> “They invite just one teacher for the training. When that person is absent during MDA or uninterested, others are left guessing what to do” (IDI, SHEP Teacher 5).
>
> “I wasn’t trained. I either had to ask my colleague what to do or try to remember back to last year, maybe that is how it’s done” (IDI, Head Teacher 3).

Use of teachers as primary drug distributors was criticized in terms of accuracy, trust, and safety. Respondents claimed that teachers are not proficient in pharmacovigilance and in managing drug reactions. One respondent said:

> “Distribution should be done by health workers rather than teachers. Parents will feel comfortable when it is health workers” (KII, NTD Program Coordinator 1).

Participant added that despite the shortage of health workers, that would not make it possible to cover all schools, even their partial involvement would raise confidence of parents, as society is more likely to trust qualified health professionals when it comes to medical interventions rather than leaving everything to teachers who are not well-trained.

### Inadequate community sensitization and engagement

Inadequacy of community-based awareness programs and engagement activities was cited as one of the main reasons for misinformation to circulate and parental resistance. The health education program was often not launched until a few days before the MDA began. This restricts the ability to effectively sensitize the community about the program.

One respondent shared that:

> “We don’t do enough community sensitization, even though we don’t effectively engage the community because money comes late… we always do things in a rush, and parents don’t get well informed.” (KII, NTD Program Coordinator 2). 1.5

This short time makes it impossible for schools to properly prepare and hold informational meetings with parents. Another respondent said that:

> “Some communities don’t even get information about the drug distribution until the morning of. We don’t even properly conduct Parent Teacher Association meetings, which creates too little confidence.” (IDI, SHEP Teacher 8).

The low and late community sensitization was a common problem experienced in both study districts. Education efforts are often not launched until a few days before the campaign hits the streets. One respondent shared that:

> “Everything comes a few days before MDA, and there is not enough time to do sensitization. That makes us rush it, and parents don’t get well informed.” (KII, NTD Program Coordinator 2).

The late educational campaign restricts the community’s knowledge and trust in the program. Short-term sensitization makes it impossible for schools to properly prepare and hold informational meetings with parents. Another respondent said that:

> “Some communities don’t even get information about the drug distribution until the morning of. That’s not enough to build confidence.” (IDI, SHEP Teacher 8).

Limited time hampers the capacity of community leaders, teachers, and health workers to provide sufficient awareness to end myths and rumors surrounding school-based preventive chemotherapy.

### Insufficient program funds

Respondents also expressed concern about inadequate funding provision from central authorities at the national, regional, or district level. There was a delay in receiving the financial resources, which hampered the organization of training, awareness campaigns, and subsequently the mass drug administration itself. As one respondent said:

> “There’s always a late release of funds…, training and all that stuff is usually done a few days before the program in a rush.” (KII, NTD Program Coordinator 1).

In some cases, lack of funds led to the skipped MDA rounds for some years, which weakened program credibility, effectiveness, and reduced community trust.

> “Sometimes we skip MDA for several years because we don’t have money to implement it…, the last time we conducted it was 2021” (IDI, SHEP Teacher 9).

Implementation of school-based preventive chemotherapy needs adequate funds to support the continuous execution of mass drug administration activities.

### Gaps in Coverage and Fidelity to Protocol

Despite the program being initially designed for annual implementation, participants acknowledged persistent gaps in frequency, duration, and coverage. Private schools were challenging to reach, often because they were not registered with the district Ghana Education Services or because their administrators declined to participate. Although the program is supposed to reach every school-age child in endemic areas, this has been far different in Gomoa East and Awutu Senya. As one district health official put it plainly:

> “We are never at 100 percent in reaching schools and students…, Coverage is always low; we have several schools that refuse to participate, especially the private schools.…, Some were not registered in the district, others are new schools that have not been incorporated into the program”. (KII, NTD Program Coordinator 1).

Out-of-school children were almost entirely excluded, despite being at high risk. Additionally, some public schools reported that pupils deliberately absented themselves on mass drug administration days due to fear, which further limited coverage.

> “Out of school children hardly come for the drug, maybe 50 out of 100,000 children; some children do not attend on a day of MDA, and some who attend were not able to take the drug because they reject… So generally, we have low coverage.” (IDI, SHEP coordinator 2).

Issues related to program duration were also evident. Some schools completed the exercise in only one or two hours, far shorter than recommended, which increased the likelihood of rushed dosing and reduced supervision of side effects. Respondents said that:

> “We do the exercise for only one day…if the child is absent, then he/she simply misses it out. There’s no follow-up, no make-up day.” (IDI, SHEP Teacher 2). “The time is too short, only a day in each school. The ones not there are missing out, and that’s how you get lower coverage.” (KII, NTD Program Coordinator 4)

Participants also noted that although the program was intended to be annual, delays in drug supply or funding sometimes caused irregular scheduling. These inconsistencies undermined the intended fidelity of the intervention.

> “The last time we did it was three years ago… there has not been stability. This program has no constant schedule, children miss treatments.” (KII, NTD Program Coordinator 1).
>
> “Skipping years diminishes an impact… transmission of schistosomiasis goes on, because these children are not treated all the time” (KII, NTD Program Coordinator 4).
>
> “If it is every year, parents and kids would get to know and prepare, but now it is coming after three years or more, for instance, the last MDA was in 2021, so when it comes again, resistance will be even more (IDI, Head Teacher 5).

Relying on school-based distribution of drugs alone cannot cover all deserving children, particularly those who are out of school, those engaged in informal labor, or who have dropped out, and pre-school-aged children. These children will still be at high risk of acquiring infection and will continue to be a potential source of schistosomiasis transmission, which could undermine programmatic objectives. To achieve higher coverage for preventive chemotherapy, there is a need to rethink additional delivery models.

### Facilitators of school-based preventive chemotherapy program for schistosomiasis

Three major themes emerged as a facilitator of school-based preventive chemotherapy program implementation in both districts: Commitment of frontline implementers and role modelling, Pre-MDA community awareness, and collaboration between health and education sectors. A detailed breakdown of themes identified is presented in Table 3.

**Table 3:**
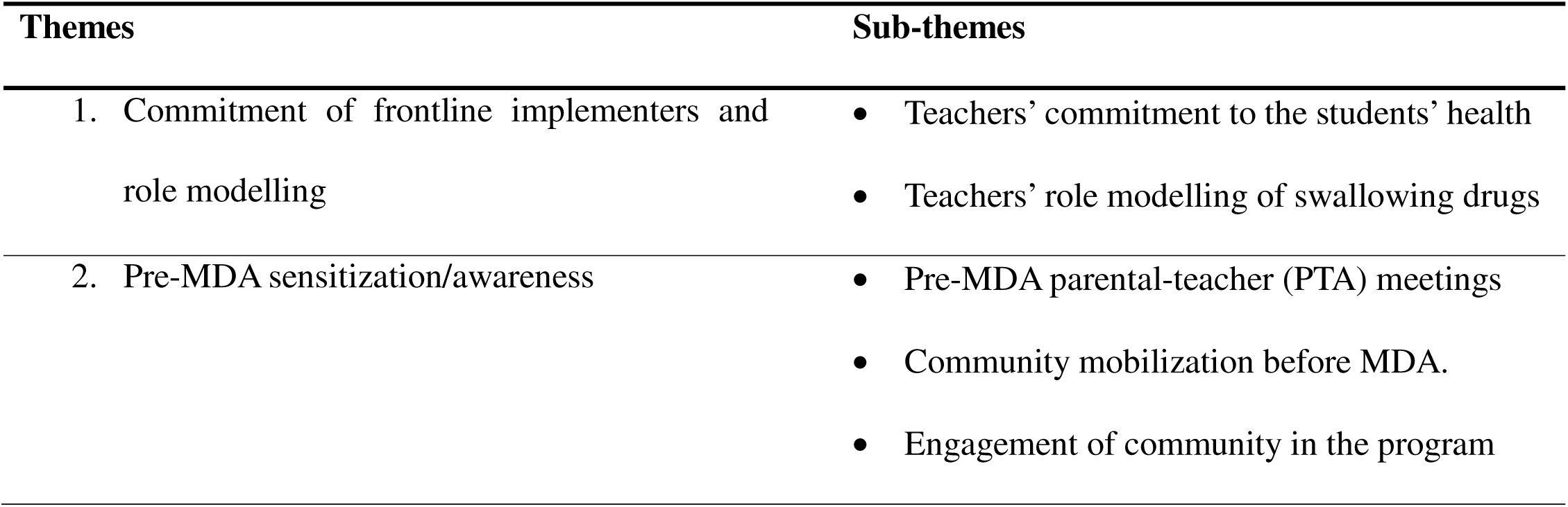
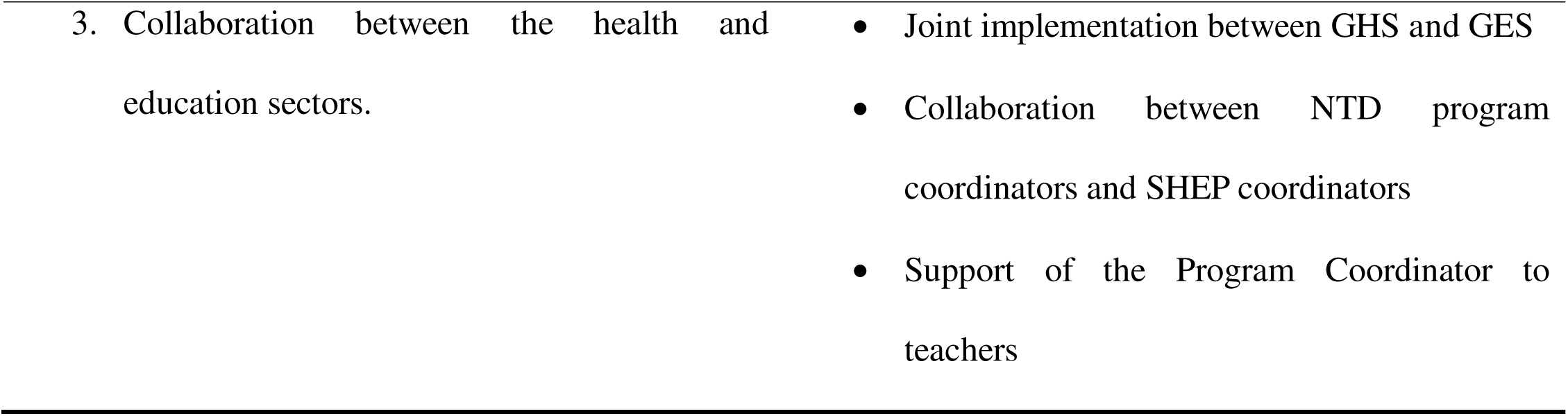
Key themes emerged as facilitators in implementing school-based preventive chemotherapy.

### Commitment of frontline implementers and role modelling

One of the strongest factors that facilitated the implementation of the school-based preventive chemotherapy program was the commitment displayed by teachers. Although teachers are not health personnel, they were central in driving mass drug administration processes to the school level. Teachers played key roles in drug logistics, mobilizing students and parents, administering drugs, supervising pupils, observing side effects, and referring those who showed reactions to health facilities. One respondent said:

> When it comes to deworming, I stop other tasks and ensure all students take the drugs. I really care about students’ health…, because I’m a victim of schisto”. (IDI, Head Teacher 1).

In schools where rumors about drugs had spread, teachers were taking drugs first in front of students as role models to demonstrate that it’s safe. and calm students down. Several participants said that children were more willing to take the drugs when they saw teachers take them first. Respondents said:

> “I always take mine first. The children watch me…, When the student sees that their teacher doesn’t suffer any side effects, they all want to be first in line for his tablets. It makes a difference.” (IDI, SHEP Teacher 8).

The role modelling and proximity of teachers to students reassured students about the safety of the drugs and hence increased their confidence in swallowing the drugs.

### Pre-MDA sensitization and awareness campaigns

Although sensitization was carried out in a rush or within a short time, it helped in reducing rumors and correcting misconceptions, while increasing the general positive attitude toward school-based preventive chemotherapy programs. One respondent said that:

> “We do explain to the parents and students before we give drugs…, we assure them that it’s safe. We move out with a microphone to announce (IDI, SHEP Teacher 9).

These communications were facilitated through Parent-Teacher Association (PTA) meetings, which provided another important means of engagement. One respondent said:

> “Mmh…, yes, we do explain the MDA program through PTA Meetings, we call the parents, that ‘we are going to start deworming your children, it is safe…. It works very much” (IDI, Head Teacher 1).

Community engagement through Teacher-parent meetings and announcements in churches and mosques helped to improve understanding. Teachers noted that sensitization was most effective when conducted regularly before the mass drug administration exercise.

### Strong coordination and collaboration between the health and education sectors

Strong collaboration between Ghana health services (GHS) and Ghana education services (GES) was widely acknowledged as an essential driver of successful school-based preventive chemotherapy delivery. District School Health Education Program (SHEP) coordinators and neglected tropical diseases (NTDs) officers worked collaboratively to plan schedules, distribute drugs, and conduct supervision. When supervision was conducted as planned, teachers felt supported and more confident. One respondent said that:

> “Anytime they come here to us from the district or regional headquarters, they do supervision and assist us in technical aspects. They ensure we get everything done, and we are correct in our documentation.” (IDI, SHEP teacher 9).

Direct support for health workers was necessary during MDA and enhanced the accuracy of program delivery. One respondent said that:

> “Sometimes nurses visit us for help to execute programs. When they come around, they guide us and solve any issue immediately.” (IDI, SHEP Teacher 6).

This support from both Ghana health services (GHS) and Ghana education services (GES) made teachers more responsible for the delivery of drugs to children. Nevertheless, this support was not comprehensive and sometimes did not exist.

## DISCUSSION

This study explored contextual barriers and facilitators influencing the implementation fidelity of school-based preventive chemotherapy (PC) for schistosomiasis in two endemic districts in Ghana. The findings demonstrate that the interaction between sociocultural beliefs, health system capacity, and organizational processes shapes implementation fidelity. Barriers such as socio-cultural beliefs, community misconceptions, drug-related fear of side effects, logistical challenges, inadequate motivation of implementers, and gaps in coverage and protocol adherence were evident; however, their influence was cumulative and mutually reinforcing, rather than independent. At the same time, facilitators, including teacher commitment, pre-MDA sensitization, and intersectoral collaboration, operated within the same system to counterbalance these constraints. This reinforces the view that fidelity is a context-dependent and system-level outcome shaped by the alignment (or misalignment) between intervention delivery and the social and institutional environment in which it is implemented.

A central finding of this study is the role of sociocultural beliefs and misconceptions as mechanisms through which both barriers and facilitators operate to influence fidelity. Myths about praziquantel, particularly associations with infertility, harm, or satanic strategies to weaken children, reflected deeper historical and sociocultural distrust of externally driven health interventions. Similar patterns have been reported in studies from Tanzania, Ethiopia, and Kenya, where negative community perceptions and low trust in preventive chemotherapy were strongly associated with reduced uptake of mass drug administration, especially when no sensitization activities were conducted (14,16,17). However, this study extends evidence by demonstrating how the timing and quality of sensitization shaped the trends of these community beliefs and misconceptions. Delayed or rushed sensitization appeared to reinforce misconceptions to spread through informal social and religious networks, ultimately creating uncertainty, reduced trust, and participation in mass drug administration and biomedical interventions. This led to over-reliance on traditional healing systems when someone got schistosomiasis or showed drug reactions. However, where early and consistent engagement occurred, the same socio-cultural structures became channels for improving acceptance, demonstrating that socio-cultural context can either undermine or reinforce fidelity depending on how it is engaged. This finding is inconsistent with (11) who found community sensitization to be effective in addressing myths and misconceptions regardless of conditions. This study identified that the impact depends on both the presence of sensitization activities and their timing, intensity, and credibility. Early and well-planned community sensitization and engagement are most important for addressing socio-cultural beliefs and misconceptions.

Drug-related fear of side effects further illustrates how biomedical realities are mediated through social processes to influence implementation fidelity. While adverse reactions to praziquantel are medically recognized (5), their visibility within school environments, combined with inadequate preparation and limited capacity to manage them, transformed these effects into social events that triggered anxiety, imitation, and widespread refusal. Consistent with studies in Liberia and Uganda, where adverse reactions to praziquantel were found to significantly affect participation, particularly when they were visible and poorly managed (18,19). However, this study provides a deeper understanding of the mechanisms through which these effects influence behavior in school settings. The clustering of side effects among students created what can be understood as a “social amplification” effect, where individual experiences triggered collective anxiety and imitation. This extends previous findings of (20) by showing that the impact of side effects may be clinical or shaped by social observation and peer influence. The study also identifies teacher role modelling as a critical counterbalancing mechanism. While previous research has acknowledged the importance of teachers in mass drug administration programs (21), this study demonstrates how their visible participation and role modelling actively normalize fears of drug reactions and rebuild confidence among students and parents, highlighting how implementers can offset biomedical concerns.

Beyond socio-cultural and side effects concerns, the findings also reveal that implementation fidelity is strongly dependent on alignment between program design and health system capacity. Logistical challenges such as inadequate measuring tools, transport barriers, and delayed funding created operational inefficiencies that interacted with human factors to weaken implementation fidelity. These logistical challenges have also been widely documented in studies across sub-Saharan Africa (17,22). However, this study advances the literature by showing how these structural constraints interact with human and social factors to produce compounded effects on fidelity. For instance, the lack of standardized measuring tools led to improvisation in height-measuring procedures, which compromised dose accuracy and undermined confidence in the intervention among implementers and the community. Similarly, delayed funding compressed the timeline for training and sensitization, which increased implementer uncertainty. Lack of training and incentives for teachers as motivation further exacerbated variability in program delivery. The reliance on teachers as primary implementers has been widely recognized as a strength of the school-based delivery model, because teachers facilitate the administration of drugs to students (23,24). However, in this study, it has shown both strengths and weaknesses. In the absence of adequate training, supervision, and incentives, their capacity to deliver the intervention consistently and manage drug reactions was limited, which undermined parents’ confidence in the school-based interventions primarily managed by teachers. However, where supportive supervision from health professionals and collaboration between the health and education sectors were present, these structural weaknesses were partially mitigated, enabling teachers to consistently adhere to protocols. This finding contributes to the literature by emphasizing that the effectiveness of school-based platforms also depends on the level of investment in implementers’ capacity and support.

The study also identified the importance of temporal and structural dimensions in shaping implementation fidelity. Short implementation windows, irregular scheduling of mass drug administration campaigns, and reliance on a single-day school-based delivery model created conditions that systematically excluded certain populations, particularly absent and out-of-school children. While previous studies have acknowledged coverage gaps in school-based mass drug administration (25,26), this study provides a more explicit explanation of how program timing and delivery structure shape these gaps. Short implementation windows, irregular scheduling, and single-day delivery models were found to systematically exclude absent and out-of-school children, which undermined both coverage and normalization of the intervention within communities. Similar challenges have been reported in studies examining the reach of school-based interventions (20,23,27). However, this study goes further by linking these structural limitations to broader issues of community acceptance and program normalization. Irregular and episodic implementation appeared to hinder the development of routine expectations around the intervention, which sustained uncertainty and resistance over time. In contrast, predictable and continuous engagement made school-based preventive chemotherapy socially acceptable, easier to operationalize, and smoother to implement. This confirms the importance of temporal consistency of delivery and routine social expectations as a determinant of operational success.

This study advances existing evidence by shifting the analytical focus from isolated barriers and facilitators to understanding how they interact within complex systems to shape implementation fidelity. While prior studies have often treated these factors as discrete determinants, this study demonstrates that their effects on program implementation fidelity are interdependent and mutually reinforcing. The findings point to the need for a fundamental reorientation of school-based preventive chemotherapy from a periodic campaign to a routine, socially embedded activity. Strengthening early and continuous community engagement through trusted local actors, ensuring alignment between program design and logistical capacity, investing in the training, motivation, and support of frontline implementers, and integrating school-based approaches with community-based strategies. There is also a need to leverage innovation to develop technology-assisted reporting and supervision systems, including mobile applications, digital dashboards, and GIS-enabled tracking tools, to improve real-time monitoring of implementation progress, coverage, and fidelity, enabling timely adjustments to program protocols.

### Strengths and limitations of the study

The study has several strengths, including the use of a qualitative design that enabled in-depth exploration of implementation processes across multiple stakeholder groups, thereby capturing the complexity of real-world program delivery. The integration of theoretical frameworks such as the Consolidated Framework for Implementation Research and the implementation fidelity framework strengthened the analytical depth and interpretation of findings, and the inclusion of both health and education sector actors allowed for triangulation of perspectives, enhancing the credibility of the results. However, the exclusion of parents and students limits the ability to fully capture demand-side perspectives. Despite this limitation, the study provides important lessons about the mechanisms shaping implementation fidelity of school-based preventive chemotherapy.

## Conclusion

The study demonstrates that implementation fidelity of school-based preventive chemotherapy (PC) for schistosomiasis is shaped by interacting sociocultural, organizational, and health system factors rather than by technical design alone. Barriers such as socio-cultural beliefs, fear of side effects, logistical constraints, and limited support to frontline implementers collectively undermine coverage, consistency in delivery, and adherence to protocol, while facilitators, such as effective sensitization, teacher commitment and role modelling, and strong collaboration between health and education sectors, enhance fidelity. Improving the school-based preventive chemotherapy programs requires context-responsive strategies, including building community trust, effective communication, and system readiness, strengthening community engagement, ensuring adequate resources and training for implementers, and enhancing coordination across sectors. It is essential to address these interconnected factors for improving implementation fidelity to maximize the impact of schistosomiasis control efforts in endemic settings.

## Data Availability

Datasets are available from the corresponding author upon reasonable request.

## Acknowledgement

The author gratefully acknowledges the:

- Ghana Health Service, the Regional and District Health Directorates, and the District Education Directorates for their administrative support.
- Mr. Kwabena Daku, Gomoa East district NTDs program coordinator, and Ms. Robin Elizabeth, Awutu Senya East NTDs program coordinator, for their cooperation throughout the study.
- Academic Supervisors from the Department of Social and Behavioral Sciences, University of Ghana, for mentorship and support provided in refining this study and manuscript development.
- WHO-TDR, the Special Program for Research and Training in Tropical Diseases, for financial support of this research under the Postgraduate Training Scheme Scholarship.

## Supplementary Files

### List of Supplementary Figures

**S1 Figure**: Conceptual framework illustrates contextual factors influencing implementation fidelity of school-based preventive chemotherapy (PC) for schistosomiasis.

**S2 Figure:** Map of the selected study area (Gomoa East and Awutu Senya East districts) in the Central Region of Ghana.

## List of Supplementary Tables

**S1 Table:** Summary of participant characteristics and distribution by role and district.

**S2 Table:** Identified barriers to implementation fidelity of school-based preventive chemotherapy for schistosomiasis.

**S3 Table:** Identified facilitators to implementation fidelity of school-based preventive chemotherapy for schistosomiasis

## List of Supplementary Appendices

**S1 Appendix:** In-Depth Interview Guide.

**S2 Appendix:** Key Informant Interview Guide.

**S3 Appendix:** Participant information sheet

**S4 Appendix:** Informed Consent Form.

**S5 Appendix:** Ethical approval letter from the Ghana Health Service Ethics Review Committee (Approval No: GHS-ERC 053/05/25).

**S6 Appendix:** Study Permission Letter from the Central Region Health Directorate

## Authors’ Contribution

– Haruna Moshi: study protocol and methods design, fund acquisition and resources mobilization, data collection and tools development, data analysis, report and manuscript writing, and editing.
– Franklin N. Glozah: study protocol and methodology refinement, Conceptualization, supervision, Data collection tools validation, report and manuscript writing, review and editing.
– Betwel J. Msugupakulya: manuscript writing, review, and editing.
– Mahnaz Vahedi: manuscript writing, review, and editing

## REFERENCES

1. Braun V. Using thematic analysis in psychology. 2006;3:77–101.

2. Damschroder LJ, Reardon CM, Widerquist MAO, Lowery J. The updated Consolidated Framework for Implementation Research based on user feedback. Implement Sci [Internet]. 2022;0:1–16. Available from: 10.1186/s13012-022-01245-0

3. Carroll C, Patterson M, Wood S, Booth A, Rick J, Balain S. A conceptual framework for implementation fidelity. Implement Sci. 2007;2(1):1–9.

4. WHO. A road map for neglected tropical disease 2021-2030 [Internet]. 2020. Available from: http://apps.who.int/bookorders.

5. WHO. WHO GUIDELINE on control and elimination of human schistosomiasis. 2022.

6. Kankpetinge C, Tekpor D, Dongdem AZ, Frimpong K, Odonkor S. Prevalence and Risk Factors of Urinary Schistosomiasis among School-Aged Children in Devego Sub-Municipal, Ketu North Municipality, Volta Region, Ghana. Int J Trop Dis Heal. 2022 Jun 11;1–10.

7. Tetteh-quarcoo PB, Forson PO, Amponsah SK, Ahenkorah J, Opintan JA, Ocloo JEY, et al. medical sciences Persistent Urogenital Schistosomiasis and Its Associated Morbidity in Endemic Communities within Southern Ghana: Suspected Praziquantel Resistance or Reinfection? 2020;

8. Opoku-Kwabi D, Sevor B, Sarpong EA, Sam PK, Frimpong AA, Marfo PA, et al. Prevalence of schistosomiasis among school children at Esuekyir community in the Central Region of Ghana. BMC Infect Dis. 2024 Dec 1;24(1).

9. Duah E, Kenu E, Adela EM, Halm HA, Agoni C, Kumi RO. Assessment of urogenital schistosomiasis among basic school children in selected communities along major rivers in the central region of ghana. Pan Afr Med J. 2021 Sep 1;40.

10. GHS. Country NTD Master Plan 2021-25. 2020.

11. Forson AO, Ayeh-Kumi PF, Mohammed AR, Kwame Sraku I, Myers-Hansen GA, Afrane YA. Knowledge, Perceptions, Challenges and opportunities in achieving sustainable coverage of mass drug administration towards the control and elimination of Schistosomiasis and Soil Transmitted Helminths in hard-to-reach communities of Ghana. PLoS Negl Trop Dis [Internet]. 2024;2024-Novem:1–13. Available from: 10.1371/journal.pntd.0012664

12. Bartlett AW, Mendes EP, Dahmash L, Palmeirim MS, de Almeida MC, Peliganga LB, et al. School-based preventive chemotherapy program for schistosomiasis and soil-transmitted helminth control in Angola: 6-year impact assessment. PLoS Negl Trop Dis [Internet]. 2023;17(5):1–19. Available from: 10.1371/journal.pntd.0010849

13. Koudessi cfe. Uptake of praziquantel mass-drug administration for schistosomiasis control among school-age children in Kpando municipality in the Volta region of Ghana by [Internet]. 2020. Available from: http://ugspace.ug.edu.gh

14. Mahende MK. Implementation fidelity of community drug distributors during preventive chemotherapy (PCT) of neglected tropical disease program in Pwani region, Tanzania. 2020;(November).

15. Adam AA. Assessing implementation fidelity of community-based integrated mass drug administration for neglected tropical disease control in Kano State, Nigeria. 2017;

16. Hungu CW. Adherence of Community Health Volunteers to Mass Drug Administration Guidelines for Schistosomiasis Control in Western Kenya. Sustain [Internet]. 2019;11(1):1–14. Available from: http://scioteca.caf.com/bitstream/handle/123456789/1091/RED2017-Eng-8ene.pdf?sequence=12&isAllowed=y%, 10.1016/j.regsciurbeco.2008.06.005%, https://www.researchgate.net/publication/305320484

17. Gebreyesus TD, Makonnen E, Tadele T, Mekete K, Gashaw H, Gerba H. Ef fi cacy and safety of praziquantel preventive chemotherapy in Schistosoma mansoni infected school children in Southern Ethiopia: A prospective cohort study. 2023;(March):1–10.

18. Agboraw E, Sosu F, Dean L, Siakeh A, Thomson R, Kollie K, et al. Factors influencing mass drug administration adherence and community drug distributor opportunity costs in Liberia: a mixed-methods approach. Parasites and Vectors [Internet]. 2021;14(1):1–11. Available from: 10.1186/s13071-021-05058-w

19. Muhumuza S, Olsen A, Katahoire A, Nuwaha F. Uptake of Preventive Treatment for Intestinal Schistosomiasis among School Children in Jinja District, Uganda: A Cross-Sectional Study. 2013;(May).

20. Torres-vitolas C, Dhanani N, Id FMF. Factors affecting the uptake of preventive chemotherapy treatment for schistosomiasis in Sub-Saharan Africa: A systematic review [Internet]. 2021. 1–33 p. Available from: 10.1371/journal.pntd.0009017

21. Sangare M, Diabate AF, Coulibaly YI, Tanapo D, Thera SO, Dolo H, et al. Understanding the barriers and facilitators related to never treatment during mass drug administration among mobile and migrant populations in Mali: a qualitative exploratory study. BMJ Glob Heal. 2024;9(10):1–15.

22. Dzidzornu D, Id O, Agbenu IA, Nyamekye MA, Appiah-agyekum N. Challenges of implementation of the preventive chemotherapy neglected tropical diseases programme in Ghana. 2023;1–13. Available from: 10.1371/journal.pntd.0011116

23. Osei FA, Newton SKT, Nyanor I, Osei-Yeboah E, Amuzu EX, Mensah NK, et al. Awareness of and participation in mass drug administration programs used for onchocerciasis control in the Atwima Nwabiagya North District, Ghana. Glob Heal Res Policy [Internet]. 2023;8(1). Available from: 10.1186/s41256-023-00331-0

24. Kiesolo FN, Sampa M, Moonga G, Michelo C, Jacobs C. uptake of the mass drug chemotherapy for schistosomiasis in a selected urban setting in. 2023;(April):1–8.

25. Campbell SJ, Osei-atweneboana MY, Id RS, Koukounari A, Cunningham L, Armoo SK, et al. The COUNTDOWN Study Protocol for Expansion of Mass Drug Administration Strategies against Schistosomiasis and Soil-Transmitted Helminthiasis in Ghana. 2018;(3).

26. Onasanya A, Bengtson M, Oladepo O, Van Engelen J, Diehl JC. Rethinking the Top-Down Approach to Schistosomiasis Control and Elimination in Sub-Saharan Africa. Front Public Heal. 2021;9(February):1–6.

27. Coulibaly JT, Hürlimann E, Patel C, Silué DK, Avenié DJ, Kouamé NA, et al. Optimizing Impl-Transmitted Helminthiasis and Intestinal Transmission of Preventive Chemotherapy against Soil Schistosomiasis Using High-Resolution Data: Field-Based Experiences from Côte d’Ivoire. Diseases. 2022;10(4):1–14.

28. Asare KK, Boampong JN, Duah NO, Afoakwah R, Sehgal R, Quashie NB. Synergism between Pfcrt and Pfmdr1 genes could account for the slow recovery of chloroquine-sensitive Plasmodium falciparum strains in Ghana after chloroquine withdrawal. J Infect Public Health [Internet]. 2017;10(1):110–9. Available from: 10.1016/j.jiph.2016.02.004

